# Assessment of pathogens in flood waters in coastal rural regions: Case study after Hurricane Michael and Florence

**DOI:** 10.1101/2022.08.16.22278858

**Authors:** Moiz Usmani, Sital Uprety, Daisuke Sano, Joanna Shisler, Avinash Unnikrishnan, Thanh H. Nguyen, Antarpreet Jutla

**Affiliations:** Environmental Engineering Sciences, University of Florida, Gainesville, FL, USA; Department of Civil and Environmental Engineering, University of Illinois at Urbana-Champaign, Urbana, IL, USA; Department of Civil and Environmental Engineering, Tohoku University, Sendai, Japan; Institute for Genomic Biology, University of Illinois at Urbana-Champaign, Urbana, IL, USA; Department of Microbiology, University of Illinois at Urbana-Champaign, Urbana, IL, USA; Civil and Environmental Engineering, Portland State University, Portland, OR, USA

## Abstract

The severity of hurricanes, and thus the associated impacts, is changing over time. One of the understudied threats from damage caused by hurricanes is the potential of cross-contamination of water bodies with pathogens in coastal agricultural regions. Using microbiological data collected after hurricanes Florence and Michael, this study shows a dichotomy in the presence of pathogens in coastal North Carolina and Florida. *Salmonella Typhimurium* was abundant in water samples collected in the regions dominated by the swine farms. A drastic decrease in *Enterococcus spp*. in Carolinas is indicative of pathogen removal with flooding waters. Except for the abundance presence of *Salmonella*, no significant changes in pathogens were observed after Hurricane Michael in the Florida panhandle. We argue that a comprehensive assessment of pathogens must be included in decision-making activities in the immediate aftermath of hurricanes to build resilience against risks of pathogenic exposure in rural agricultural and human populations in vulnerable locations.

## Introduction

Hurricanes cause significant economic damage (1), traditionally measured in terms of loss of civil infrastructure, primarily in urban locations in the United States. The vulnerability of highly dense metropolitan coastal communities to hurricanes under both current and changing climates is well-documented (2,3). However, an understanding of the susceptibility of rural, agricultural-dominant, human communities to the short- and long-term impacts of hurricanes is still lacking (4). Rural regions tend to receive limited financial resources and government assistance, which may decrease the resilience of these communities to recover from the effect of extreme natural events (4). In livestock-dominant agricultural communities, hurricane-induced flooding poses the risk of contaminating natural water sources with run-off from lagoons and barnyards containing animal fecal material. In turn, this could increase the exposure of pathogens to humans. A handful of studies have highlighted the role of pathogens (5) in agricultural regions after hurricanes, despite the ubiquitous presence in rural communities. In 2018, there was concern that water from Hurricane Michael may overtop wastewater treatment plants and sanitary sewer systems, which may lead to contamination of drinking water supply lines (6)(7).

In 2018, the Atlantic Hurricane season was dominated by hurricanes Florence and Michael. Hurricane Florence made landfall near Wrightsville Beach, NC, as Category 1 storm on September 14, 2019 (8). Despite the storm’s relatively low wind speeds, Hurricane Florence wreaked havoc with towering storm surges and historical rainfall (8)(9). Less than four weeks later, Hurricane Michael made landfall near Mexico Beach, FL, as a Category 4 hurricane (10). Both hurricanes were destructive, but the means by which each storm caused the damage was unique. Hurricane Michael produced only a fraction of the rainfall that deluged North Carolina (11) compared with hurricane Florence in the Florida panhandle.

The lack of adequate information on the presence and prevalence of pathogens of clinical importance after hurricanes is the key motivation for the current study. Therefore, the objective here was to provide a survey assessment for the genes from key bacterial pathogens after hurricane-induced flooding in the rural coastal locations of North Carolina and Florida. Our anticipated goal is to initiate the dialogue through the characterization of the vulnerability of humans in terms of exposure to clinically significant pathogens after hurricanes.

## Materials and Methods

### Selection of Sampling Locations

The first step toward accurately identifying sampling locations was to characterize the flooded regions after hurricanes. Inundation resulting from extreme rainfall can be modeled using traditional hydrological and hydrodynamic models. However, here we used a relatively new method for mapping inundation that is based on geomorphometric principles of the landform (12–14). The basic premise is to let topography dictate how water will fill a particular landscape. It allows a fast but static computation and accurate identification of locations likely to be inundated after heavy rainfall. Digital elevation model (DEM) data at 30 m resolution was used to spatially capture the locations and estimate the inundation depths due to any known amount of rainfall. Details on inundation mapping are provided in previously published work (15). Sampling locations for North Carolina and Florida were selected by mapping the flood extents of each hurricane, identifying agriculture or wastewater infrastructure exposed to flooding, and then selecting accessible water bodies downstream of the point of interest. Twenty-six locations were sampled in North Carolina on October 7, 2018 (3 weeks post-hurricane), ranging from the coast to about 100 miles inland. In the Florida panhandle, 11 locations were sampled on October 27, 2018 (2 weeks post-hurricane), with the locations tailored to investigate the impacts of storm surge on pathogen transport in coastal communities and flooded water treatment facilities, covering a geographical extent of Pensacola to Port St. Joe, Florida.

The locations of swine farms were obtained from the North Carolina Department of Environmental Quality (NCDEQ) Animal Feeding Operations Program (16). The 2283 permitted farm locations were overlaid onto the flood map, which was then queried by flood depth. All swine farms with flood depths of five feet or greater were selected as potential sampling sites because there was high certainty about flooding at the location, resulting in the selection of 40 swine farms. This number is similar to the number arrived at by the post-hurricane report given by the NCDEQ, in which 28 swine facilities reported lagoon discharging. An additional eight reported inundation (surface water surrounding and flowing into the lagoon), and eight more reported that lagoons were at full capacity and likely to overtop (17). Similarly, unflooded farms were identified, and 23 locations were selected from the list of unflooded farms by visual inspection to meet two criteria. First, the unflooded location must be relatively close to flooded farms of interest, making travel and collection if samples less burdensome. Second, the flood maps must show the location as free of flooding to a high certainty (i.e., not located on or near the boundary of the flood extent). While in the field, water samples were taken at publicly accessible locations downstream and near the selected farms. The sampling locations are shown in Figures 1 and 2.

**Figure 1:**
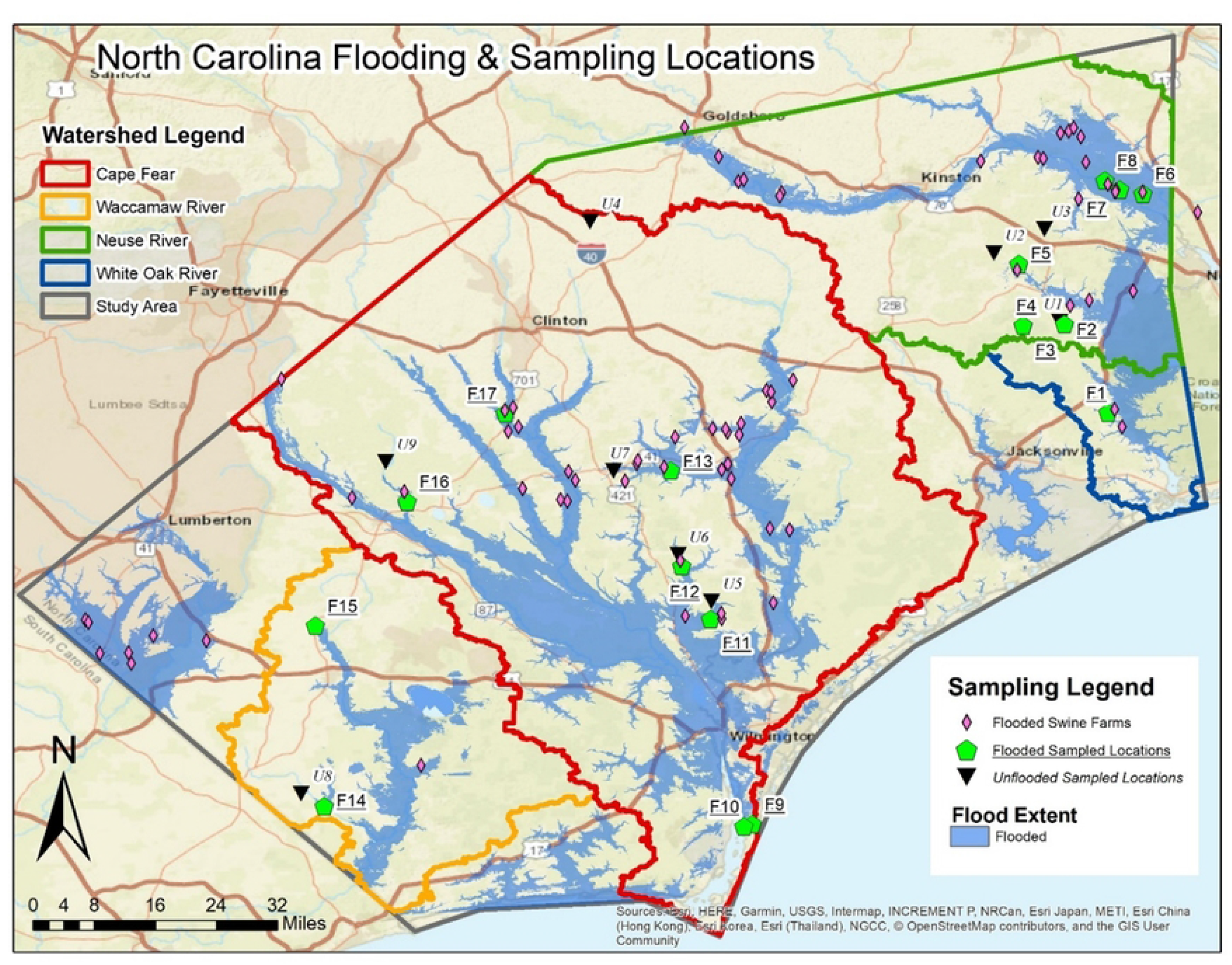
North Carolina Flooding and Sampling Locations

**Figure 2:**
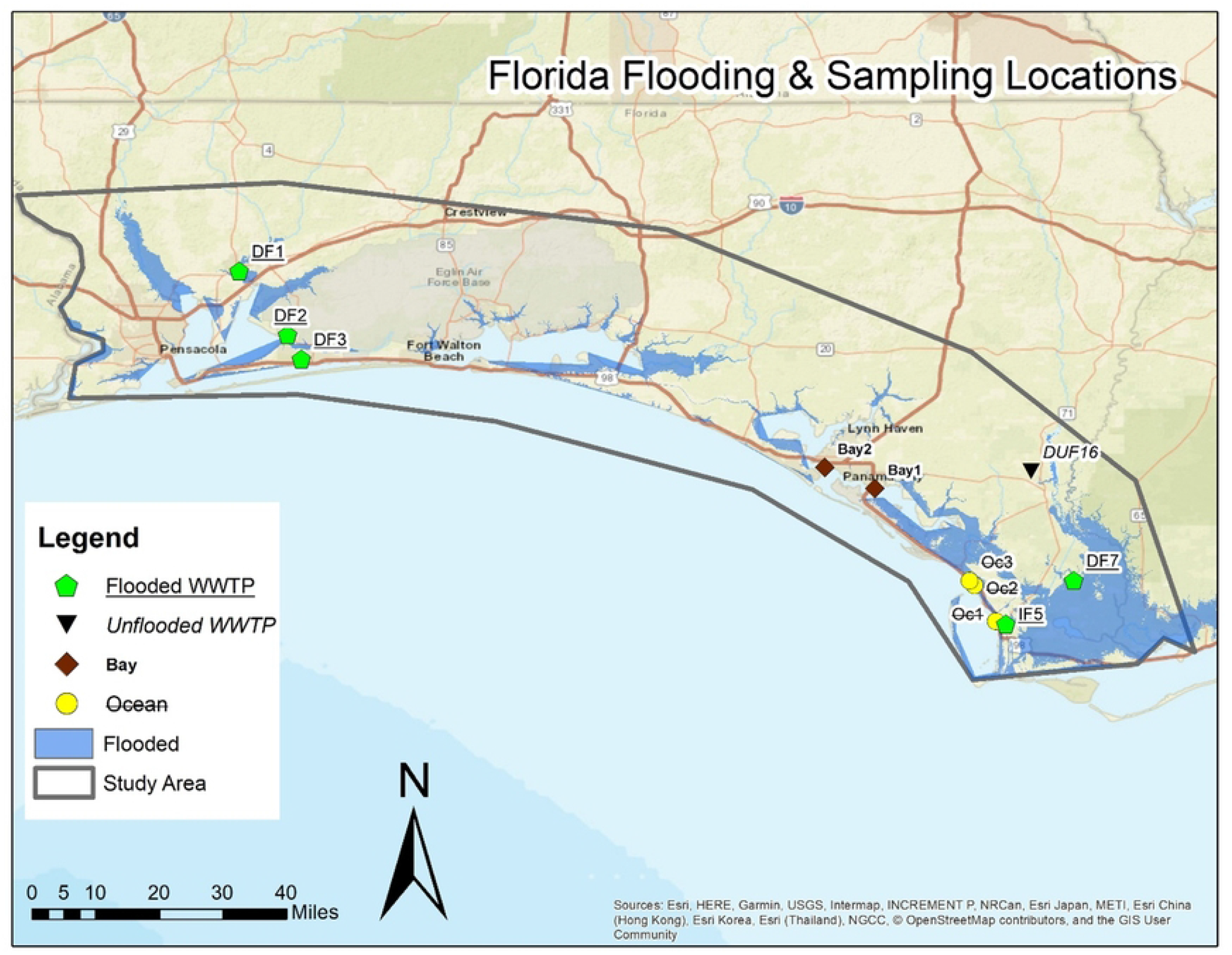
Florida Flooding and Sampling Locations. DF = Domestic, Flooded WWTP. DUF= Domestic, Unflooded WWTP

Compared to Hurricane Florence’s effects in North Carolina, Hurricane Michael did not trigger inland flooding to the same depth or extent as observed in the Florida Panhandle. This, combined with the absence of a singular dominating industry at flood risk comparable to swine farming in North Carolina, led to an eclectic array of sampling locations in Florida. Furthermore, widespread road closures forced many sampling locations to be selected in the field based on accessibility. Water samples were taken downstream near five wastewater treatment plants (WWTP)(18), three in the ocean between Port St. Joe and Mexico Beach, and two in the bays surrounding Panama City, FL as seen in Figure 2.

### Lab testing Procedure

We sampled water in N.C. and F.L. in October 2018, post-Hurricane Florence and Michael, respectively. As with our previous fieldwork (19,20), we used sterilized plastic bags to collect and store four to eight liters of water from each source. Bags were kept in a cooler with ice packs and transferred within 24 hours to a 4°C cold room in our lab. The volume of sampled water was tracked when samples were filtered. Water samples were coagulated with 25 mM MgCl_2_ for 30 minutes before settling. Subsequently, they vacuum-filtered sequentially through a glass fiber filter with a 1.6 µm pore size (Fisher Scientific, Hampton, NH) to collect and concentrate bacteria. Each filter was cut into four equal parts. Because of different turbidities, different numbers of filters were required for water samples. A quarter of 1.6 µm filter of all filters used for a given water sample were subjected to DNA extraction by MPI FastDNA Kit for Soil Extraction (M.P. Biomedicals, Santa Clara, CA). The rest of the filters were kept for other ongoing analyses.

Quantitative polymerase chain reaction (qPCR) assays were adapted from previous studies to detect *Enterococcus spp*., two genes of *E. Coli*, Enteropathogenic *E. Coli*, Shiga-toxin producing *E. Coli Shigella* spp, *Shigella flexneri*, two genes of *Campylobacter jejuni, Campylobacter lari*, two genes of *Salmonella, Clostridium perfringens, Listeria monocytogenes*, two genes of *Vibrio cholerae, Mycobacterium* spp., *Pseudomonas* spp., *Legionella* and *Giardia lamblia* (15,21–31). Forward and reverse primers for all assays were obtained as Custom DNA Oligos (Integrated DNA Technologies, Coralville, IA). Probes were obtained from the Universal Probe Library (UPL) (Roche, Basel, Switzerland) and were labeled with 6-FAM at the 5’ end and a dark quencher dye at the 3’ end and contained a short sequence (8-9 nucleotides) of locked nucleic acids (32). Standards were obtained as gBlock Gene Fragments (Integrated DNA Technologies). Standard curves were generated by qPCR using serial dilutions (2 × 10^0^ to 2 × 10^6^ copies/μl) of a standard pool containing 24 DNA standards to validate the assays prior to use in MFQPCR. PCR inhibition was evaluated for the STA and MFQPCR analysis by including *Pseudogulbenkiania* NH8B as an internal amplification control in all environmental sample extracts and nuclease-free water. Prior to enumeration by mfqPCR, all DNA samples and standard pool dilutions underwent standard target amplification (STA) PCR to increase template DNA yields. Standard pool dilutions (2 × 10^0^ to 2 × 10^6^ copies/μl) amplified in the 14-cycle STA were used to generate standard curves for MFQPCR. 20X assays (18 μM of each primer and 5 μM probe) were pooled using 1 μl per assay and 179 μl of DNA Suspension Buffer (Teknova, Hollister, CA) to make a 0.2X TaqMan primer-probe mix. The reaction (5 μl) contained 2.5 μl 2X TaqMan PreAmp Master Mix (Thermo Fisher), 0.5 μl 0.2X TaqMan primer-probe mix, and 1.25 μl of template DNA. The PCR plate was processed with the following thermal cycle on an M.J. Research Tetrad thermal cycler (M.J. Research, Waltham, MA): 95°C for 10 min and 14 cycles of 95°C for 15 sec and 60°C for 4 min. The STA products were diluted 25-fold with100 μl of T.E. buffer and were used for mfqPCR. The sample premix (5 μl) contained 2.5 μl 2X TaqMan Master Mix, 0.25 μl 20X Gene Expression Sample Loading Reagent (Fluidigm, South San Francisco, CA), and 2.25 μl 25-fold diluted STA product. The assay mix (5 μl) contained 2.5 μl 2X Assay Loading Reagent (Fluidigm) and 2.25 μl 20X TaqMan primer-probe mix. Aliquots (5 μl) of each sample and duplicates of each assay were loaded onto a 48.48 chip (Fluidigm). mfqPCR was performed in a Biomark HD Real-Time PCR (Fluidigm) using the following thermal conditions: 70°C for 30 min, 25°C for 10 min, 95°C for 1 min, followed by 35 cycles of 96°C for 5 sec and 60°C for 20 sec.

Quantification cycle (C_q_) values and standard pool dilutions (log copies/μl) were used to generate standard curves for each assay. C_q_ values were determined by Real-Time PCR Analysis software (Fluidigm) and MFQPCR. Linear regression analysis was performed to fit the standard curves and calculate the goodness of fit (R^2^). Assay efficiencies were calculated based on the slopes of the standard curves for each MFQPCR assay to validate adequate target amplification (33). Standard curves were accepted as quantifiable if the efficiency achieved was greater than or equal to 90% and if the lower detection limit was less than or equal to 30 copies/μl. Consistent detection of NH8B throughout multiple assays indicates insignificant inhibition for qPCR amplification. The concentrations of detected bacterial genes were reported in the unit of gene copies per L of water sample.

## Results and Discussion

Water samples collected from creeks, ponds, and rivers immediately after hurricanes were tested for the presence of six common pathogenic genes [*Enterococcus spp*., *Legionella pneumophila, Mycobacteria* (atpE), Pseudomonas (gyrB), *Salmonella Typhimurium* (ttrC), *E. coli* (eaeA, uidA, ftsZ)] from 26 different sampling locations in North Carolina and Florida (Figure 3). *Enterococcus spp*. (Fig 3a), *Mycobacteria* (Fig 3c) and *Salmonella Typhimurium* (Fig 3e) were detected in almost all the flooded locations in high concentrations relative to the unflooded ones in North Carolina. *Legionella* (Fig 3b), *Pseudomonas* (Fig 3d), and *E. coli* (Fig 3f) were present in a few of the flooded locations in higher concentrations than the unflooded sampling points. Unlike North Carolina, there was no clear distinction in the detection of pathogens in the water systems in Florida after Hurricane Florence. *Enterococcus spp* (Fig 4a), *Salmonella Typhimurium* (Fig 4e), and *E. coli* (Fig 4f) were the most prevalent pathogens among the eleven sampling locations (see map in Figure 2). Presence-wise, all flooded and unflooded locations had *Salmonella Typhimurium* in their water system.

**Figure 3:**
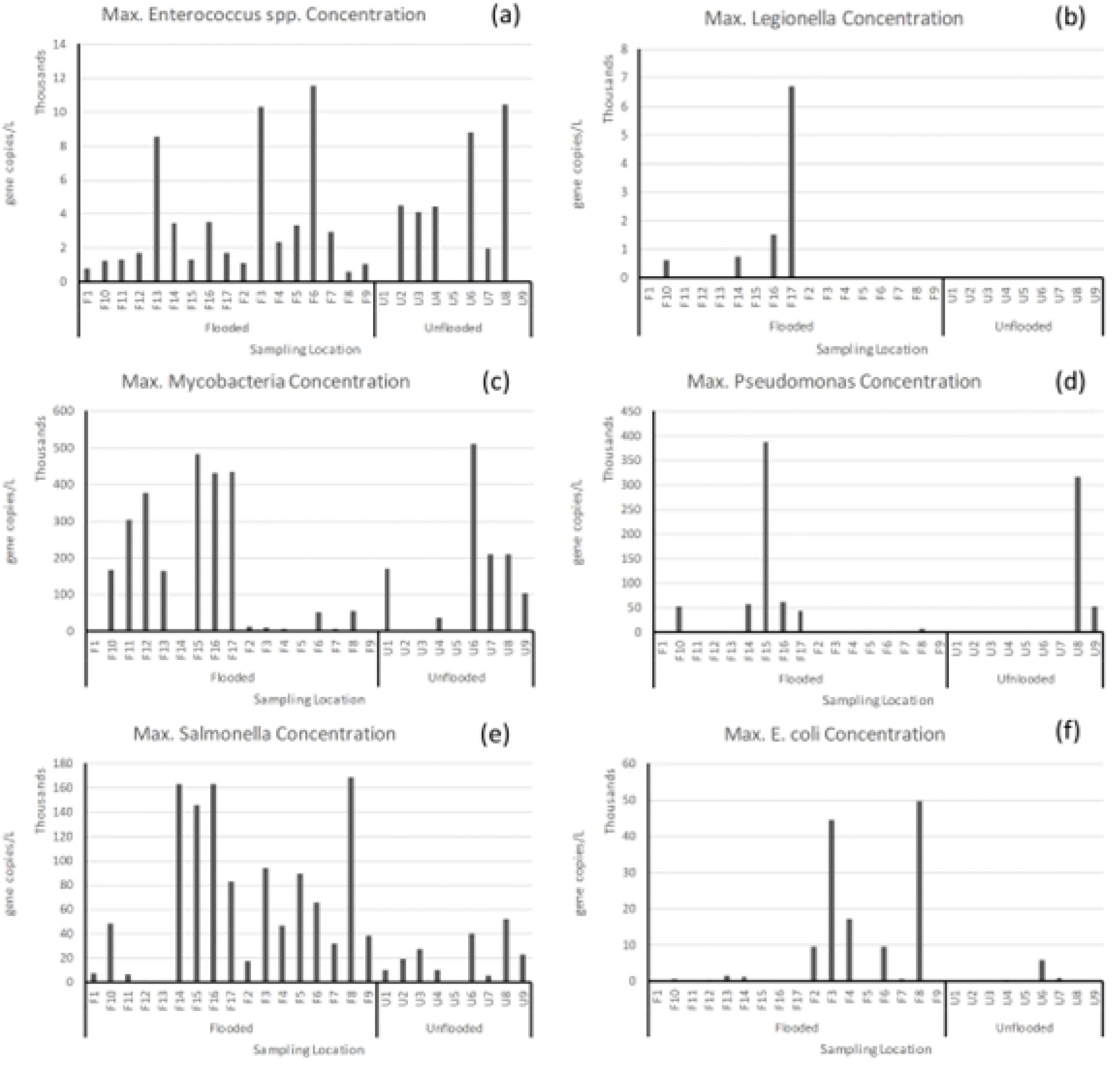
North Carolina Max. Pathogen Concentrations. **(a)** Enterococcus spp. **(b)** Legionella pneumophila, **(c)** Mycobacteria (atpE), **(d)** Pseudomonas (gyrB), **(e)** Salmonella Typhimurium (ttrC), **(f)** E. coli (eaeA, uidA, ftsZ) [F1, F2… F12 implies flooded location; U1 to U9 are unflooded sampling point)

**Figure 4:**
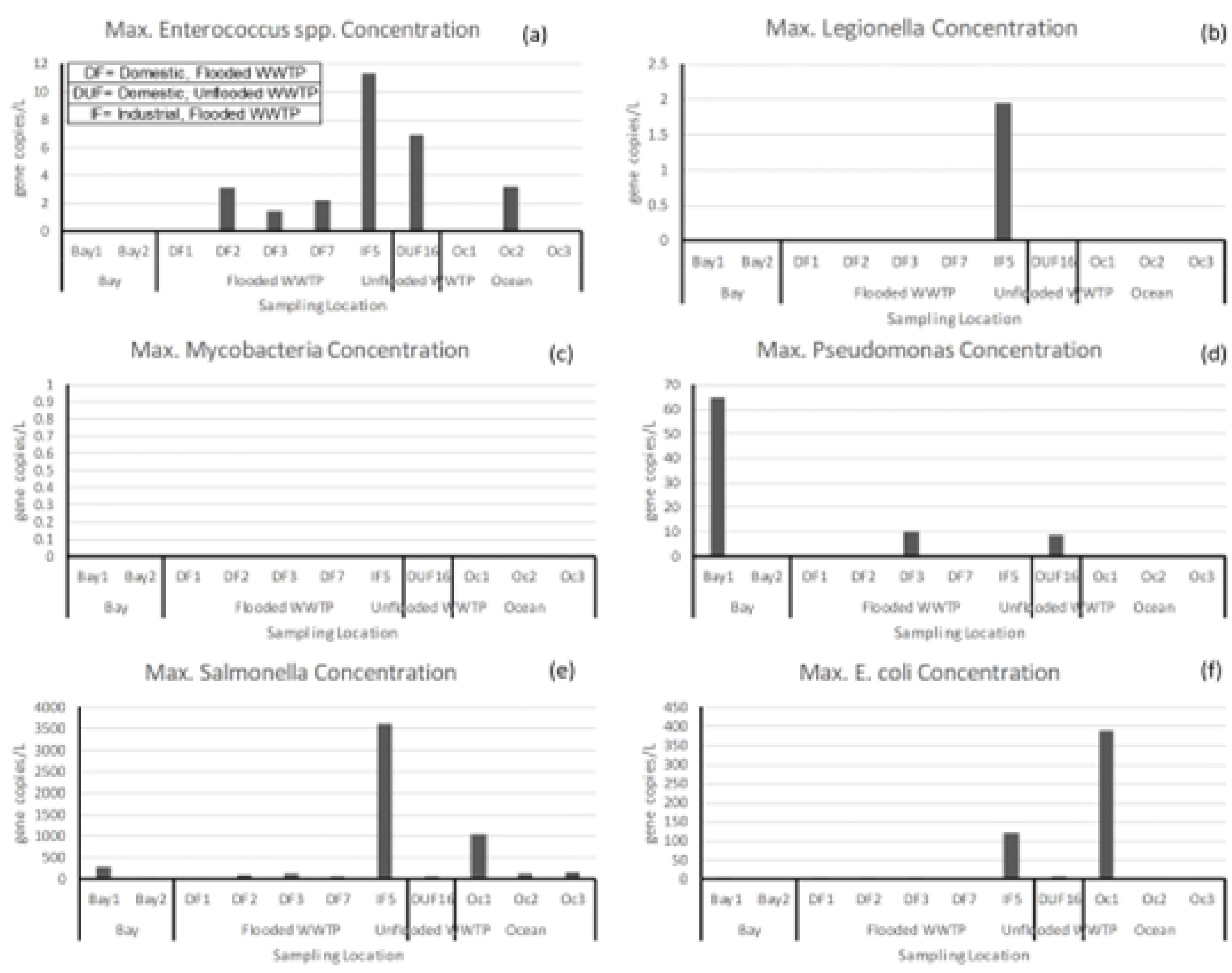
Maximum concentrations of pathogens in Florida. **(a)** *Enterococcus spp*. **(b)** *Legionella pneumophila*, **(c)** Mycobacteria (atpE), **(d)** Pseudomonas (gyrB), **(e)** *Salmonella typhimurium* (ttrC), **(f)** *E. coli* (eaeA, uidA, ftsZ) [DF = Domestic, Flooded WWTP. DUF= Domestic, Unflooded WWTP; WWTP=waster water treatment plant]

In order to explore the impacts of floods on pathogens, a probability of exceedance analysis was performed on the samples collected from North Carolina. There appears to be no difference in the presence of *Mycobacteria* with inundation (Figure 5a), as the probability of non-exceedance for flooded and unflooded samples was very similar. However, there was a marked difference in the presence and detection of *Salmonella Typhimurium*(Figure 5b) in the rural agricultural regions. Flooding appears to have strengthened the abundance of *Salmonella* when the probability of non-exceedance is greater than 50%. On further examination, the odd ratio analysis suggested that the presence of *Salmonella Typhimurium* in surface water bodies increased by 2.3 times during flooding. The increased likelihood of *Salmonella Typhimurium* during flooding may be attributable to cross-contamination of litter and associated swine activities, including run-off water from livestock farms. However, additional experiments are required to ascertain this observation fully. One of the interesting findings is with the *Enterococcus spp*. (Figure 5c) where the abundance of pathogen decreased during flooding, especially when the probability of exceedance increased to 0.5. It is plausible to the supposition that flooding water washed off the pathogen from its natural environment to coastal waters. This, therefore, decreased its concentration in the terrestrial surface water bodies. The odds ratio analysis suggested a three-fold decrease in this pathogen during floods following hurricanes.

**Figure 5:**
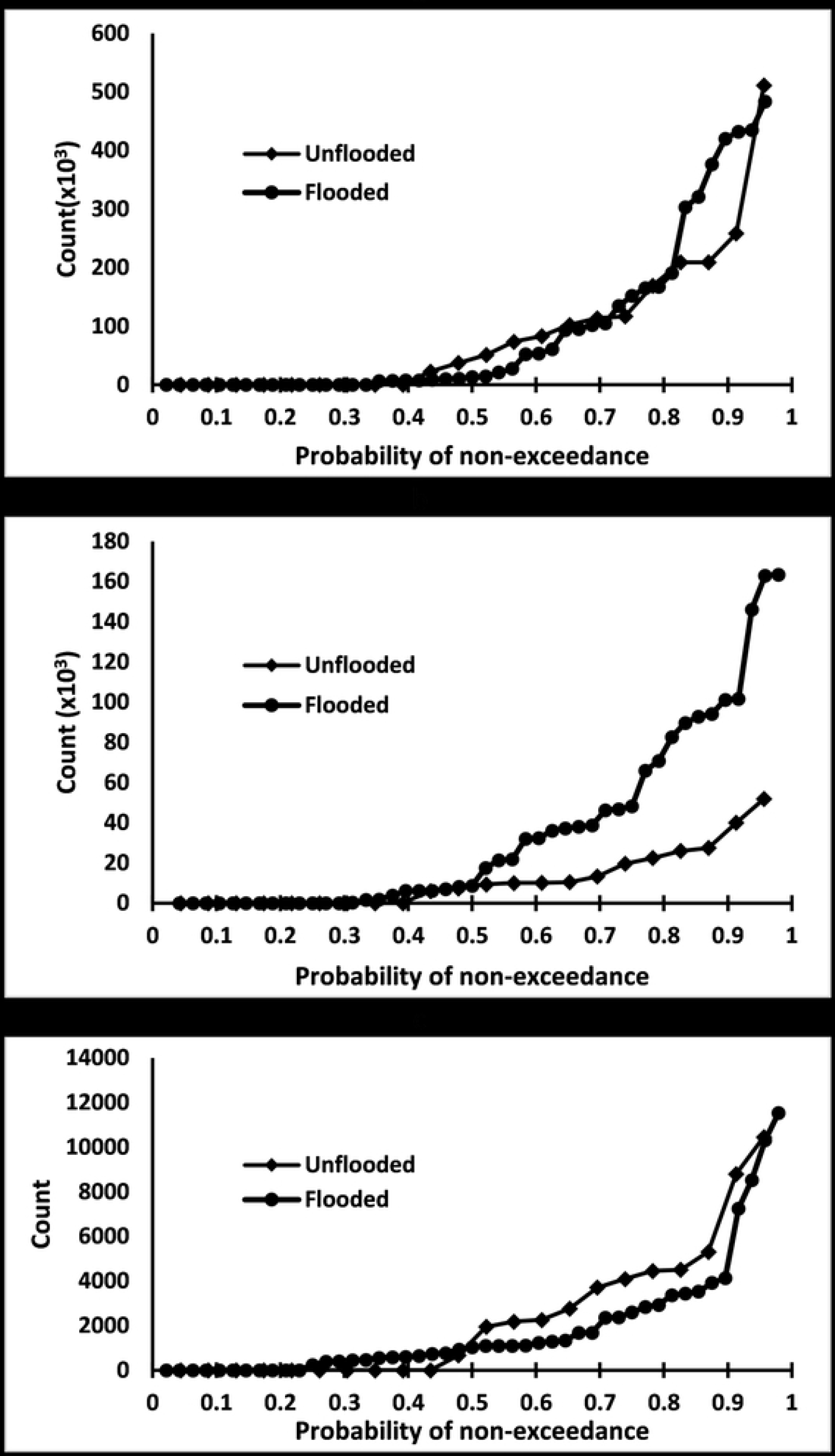
Probability of exceedance analysis for the samples collected from North Carolina. (a) *Mycobacteria*, (b) *Salmonella Typhimurium* and (c) *Enterococcus spp*.

A comparison between hurricane Florence and Michael is provided in Table 1. Hurricane Florence was characterized by record-breaking rainfall and inland flooding, whereas the damage caused by Hurricane Michael was attributed primarily to coastal (storm surge) flooding and wind damage. North Carolina contains a massive agriculture industry dominated by swine farming, while Florida lacks a singular industry with obvious potential for introducing pathogens into surface waters. These two dissimilarities are evident in the remarkable differences in pathogen concentrations between the two study areas.

**Table 1:** Hurricanes Florence and Michael Summary

| **2018 Atlantic Hurricane** | **Florence<sup>1</sup>** | **Michael<sup>3</sup>** |
| --- | --- | --- |
| *Landfall Date* | September 14 | October 10 |
| *Landfall Site* | Wrightsville Beach, NC | Mexico Beach, FL |
| *Category at Landfall* | 1 | 4 |
| *Rainfall* | 20-30 inches, 35 inches + local | < 7 inches |
| *Storm Surge* | 9 – 13 feet | 9 – 14 feet |
| *Max. Wind* | 106 mph, less severe | 155 mph, severe |
| *Inland Flooding* | Severe, 28 flood records broken<sup>2</sup> | Limited/minor |
| *Study Area* | Coastal and Sub-coastal NC | Coast of Florida panhandle |
| *Dominant Local Industry* | Swine farming<sup>4</sup> | N/A |

1, *Historic Hurricane Florence, September 12-15, 2018*. September 2018. National Oceanic and Atmosphere Administration. web. 28 February 2019. <https://www.weather.gov/mhx/Florence2018>.
2, *USGS: Florence set at least 28 flood records in Carolinas*. 13 November 2018. web. 28 February 2019. <[https://www.usgs.gov/news/usgs-](https://www.usgs.gov/news/usgs-florence-set-least-28-flood-records-carolinas) [florence-set-least-28-flood-records-carolinas](https://www.usgs.gov/news/usgs-florence-set-least-28-flood-records-carolinas)>.
3, *Catastrophic Hurricane Michael Strikes Florida Panhandle October 10, 2018*. October 2018. web. 3 March 2019. https://www.weather.gov/tae/20181010_Michael.
4, National Agriculture Statistics Service, Agriculture Statistics Board, United States Department of Agriculture. "Quarterly Hogs and Pigs." Quarterly Report. December 20, 2018. web. 4 March 2019. <[https://downloads.usda.library.cornell.edu/usda-](https://downloads.usda.library.cornell.edu/usda-esmis/files/rj430453j/bc386p647/rf555zc904/hgpg1218.pdf) [esmis/files/rj430453j/bc386p647/rf555zc904/hgpg1218.pdf](https://downloads.usda.library.cornell.edu/usda-esmis/files/rj430453j/bc386p647/rf555zc904/hgpg1218.pdf)>.

## Conclusion

Traditionally, the studies of pathogens in floodwaters are generally reported in regions with poor water and sanitation infrastructure with known knowledge of the emergence of microbes after heavy rainfall. In the continental United States, speculative assessment of pathogens is conducted after floods from the standpoint of risk of diseases in the urban human population. Perhaps this study is one of the first that has tried to shed insights on the pathogenic dangers of hurricane-induced flooding in the rural agricultural region of the U.S. The agricultural livestock regions of the U.S. are at constant threat of changes in climatic patterns, and thus effective policies should be made to safeguard these commodities. A significant portion of U.S. extensive livestock agriculture is located within a few hundred miles of the eastern coast (e.g., swine farms in N.C.). An increased occurrence of extreme events is likely to devastate the rural economies. Therefore, the significant implications of this study include an ambitious plan to develop a database for threat assessment of pathogens in the immediate aftermath of the hurricanes. We have shown that the impacts of the two hurricanes along two prominent U.S. coasts have very different pathogens. Hence, the warnings may include information on microbes that may be prevalent in the water system after the extreme events. A well-planned infrastructure plan should be in place to safeguard agricultural commodities so that pathogen spillover should be contained or anticipated in advance.

## Data Availability

All relevant data are within the manuscript.

